# Detection of fecal coliforms and SARS-CoV-2 RNA in sewage and recreational waters in the Ecuadorian Coast: a call for improving water quality regulation

**DOI:** 10.1101/2022.01.04.22268771

**Authors:** Maritza Cárdenas-Calle, Leandro Patiño, Beatríz Pernia, Roberto Erazo, Carlos Muñoz, Magaly Valencia-Avellan, Mariana Lozada, Mary Regato-Arrata, Miguel Barrera, Segundo Aquino, Stalyn Moyano, Stefania Fuentes, Javier Duque, Luis Velazquez-Araque, Bertha Carpio, Carlos Méndez-Roman, Carlos Calle, Guillermo Cárdenas, David Guizado-Herrera, Clara Lucía Tello, Veronica Bravo-Basantes, Josué Zambranod, Jhannelle Francis, Miguel Uyaguari

**Affiliations:** Red Interinstitucional para el Estudio de Ecosistemas Acuáticos de Ecuador, Guayaquil, Guayas, Ecuador; Ambiente Sociedad & Empresa Research Group. University of Guayaquil, Guayaquil, Guayas, Ecuador; Faculty of Chemical Engineering, University of Guayaquil, Cdla. Universitaria “Universidad de Guayaquil”, Guayaquil 090510, Guayas, Ecuador; Dirección Técnica de Investigación, Desarrollo e Innovación, Instituto Nacional de Investigación en Salud Pública Dr. Leopoldo Izquieta Pérez, Guayaquil, Guayas, Ecuador; Instituto de Investigaciones de Recursos Naturales, Facultad de Ciencias Naturales, Universidad de Guayaquil, Av. Raúl Gómez Lince s/n y Av. Juan Tanca Marengo, Guayaquil, Guayas, Ecuador; Laboratorio de Microbiología Ambiental, Instituto de Biología de Organismos Marinos, CONICET, Puerto Madryn, Chubut,Argentina; Centro de Referencia Nacional de Virus Exantemáticos, Gastroentericos y transmitidos por vectores. Instituto Nacional de Investigación en Salud Pública Dr. Leopoldo Izquieta Pérez, Guayaquil, Guayas, Ecuador; Dirección de Medio Ambiente-Gobierno Autónomo Descentralizado Provincial de Santa Elena, Santa Elena, Ecuador; Área Nacional de Recreación Playas Villamil, Ministerio del Ambiente y Agua, Playas, Ecuador; LAB CESTTA, Vía a Daule km 1,5. Parque California 2, bodega C 36, Guayaquil, Ecuador; Department of Microbiology, University of Manitoba, Winnipeg, Manitoba, R3T 2N2, Canada

**Author notes:** Corresponding Author: Miguel Uyaguari, 45 Chancellors Circle, Room 414D/Laboratory 118, Buller building, Winnipeg, Manitoba, R3T 2N2 Canada. Equal contribution (Maritza Cárdenas-Calle, Leandro Patiño).

**Keywords:** SARS-CoV-2, fecal coliforms, seawater, estuaries, water quality

## Abstract

Wastewater surveillance represents an alternative approach for the diagnosis and early detection of infectious agents of public health importance. This study aimed to evaluate SARS-CoV-2 and other quality markers in oxidation lagoons, estuarine areas and seawater at Guayas and Santa Elena in Ecuador. Sample collections were conducted twice at 42 coastal sites and 2 oxidation lagoons during dry and rainy seasons (2020-2021). Physico-chemical and microbiological parameters were evaluated to determine organic pollution. Quantitative reverse transcription PCR was conducted to detect SARS-CoV-2. Results showed high levels of *Escherichia coli* and low dissolved oxygen concentrations. SARS-CoV-2 was detected in sea-waters and estuaries with salinity levels between 34.2-36.4 PSU and 28.8 °C-31.3 °C. High amounts of fecal coliforms were detected and correlated with the SARS-CoV-2 shedding. We recommend to decentralized autonomous governments in developing countries such as Ecuador to implement corrective actions and establish medium-term mechanisms to minimize a potential contamination route.

**HIGHLIGHTS:**

- SARS-CoV-2 RNA was detected in estuaries, bays and the wastewater treatment systems in Playas and Santa Elena.
- High levels of fecal coliforms were detected along shorelines.
- Water quality parameters revealed a negative impact on the beaches studied associated with human activities.

## Introduction

Recreational water includes rivers, lakes and coastal waters used generally for swimming, surfing, whitewater sports, diving, boating and fishing. These waters are usually located near urban areas, where microbial and chemical contamination derived from anthropogenic activities can cause detrimental effects to human health and ecosystems (Edokpayi et al., 2017; Rodrigues and Cunha, 2017; Yuan et al., 2019; WHO, 2003). Fecal contamination and introduction of associated human pathogens (bacteria and viruses) are one of the main issues associated with recreational water use. This may occur through point source discharges (e.g. discharges of treated sewage/wastewater), runoff from urban or agricultural areas, bather and animal excreta (Bosch et al., 2005). Targeted epidemiological studies in recreational water have found a cause– effect relationship between fecal pollution and enteric or acute febrile respiratory illnesses, each of which accounts for more than 120 million and 50 million cases yearly, respectively (Shuval, 2003). Surveillance data from the United States of America (USA) during 2009 and 2010, showed 24 recreational water disease outbreaks associated with the use of natural waters, 13 of which were attributed to human pathogens. According to the reports, most of the microbial agents responsible for the outbreak included *Campylobacter jejuni, E. coli* O *Giardia intestinalis* 157:H7, *Shigella sonnei, Cryptosporidium* spp., *Giardia intestinalis, Avian schistosomes*, and *Norovirus*.

Monitoring of recreational water for fecal contamination is a common policy in developed countries. It is conducted using certain bacterial species as indicators (Paruch and Mæhlum 2012; Ramos-Ortega et al., 2008), which include total and thermotolerant fecal coliforms. These bacteria are not postulated as the causative agents of illnesses in bathers, but appear to behave similarly to the actual fecal-derived pathogens (Prüss, 1998). *E. coli*, in particular, has been proposed as one of the best indicators for gastroenteritis and dermal symptoms caused by seawater bathing (Wu et al., 2011), given its high frequency in domestic drains and longer survival time in seawater with respect to other coliforms. Other non-pathogenic fecal microorganisms such as intestinal enterococci are now being recommended to assess health risk in recreational waters (WHO, 2021). Fecal coliforms are normally monitored in beaches to detect impacts from sewage (Abdelzaher et al., 2013). Sewage sources in beaches include cesspools, septic tanks, and leaking sanitary sewer systems (Brandão et al., 2020).

The pandemic “Severe Acute Respiratory Syndrome Coronavirus 2 (SARS-CoV-2)” causing COVID-19 disease is also excreted in feces (Chen et al., 2020). Its main route of transmission occurs via respiratory droplets and human contact, and while environmental transmission has been suggested, this is still unclear (Al Huraimel et al., 2020). The virus nucleic acid has been detected in untreated and treated sewage in several countries including Australia, China, the USA, the Netherlands, France, Spain, and Italy but also in rivers impacted by urban wastewater in Japan and Ecuador (Haramoto et al., 2020; Guerrero-Latorre et al., 2020). Within this context, there are concerns that transmission from wastewater might occur or that viable viruses from its effluents reach water habitats posing a risk for the user or for the aquatic fauna. However, and based on indirect evidence, the risk of infection with SARS-CoV-2 from wastewater and recreational water bodies is considered low according to the Centers of Disease Control and Prevention (CDC, 2020a; 2020b), as virus exposure to environmental conditions would reduce its viability. A laboratory assay using the coronavirus murine hepatitis virus spiked-in raw wastewater has shown a 90% reduction of infectivity at 25 °C after 13 h but longer infectivity rate at 10 °C after 36 h. Using the same assays, the SARS-CoV-1, genetically closer to SARS-CoV-2, has been found to be viable up to 14 days at 4°C and 2-3 days at ∼20°C in raw sewage (Wang et al., 2005; Ye et al. 2016). In fact, SARS-CoV-2 was found to be infectious after 2.3 days at 20 °C and 3.8 days at 4 °C in river water, and 1.1 days at 20 °C and 2.2 days at 4 °C in seawater (Sala-Comorrera et al. 2021). While the risk of transmission of SARS-CoV-2 from water bodies needs further evidence, there are growing recommendations for its surveillance as an early warning tool for predicting outbreaks, occurrence, prevalence, and potential public health risks in the communities (Kitajima et al., 2020; Mordecai and Hewson 2020).

Monitoring of fecal contamination in recreational waters is not usually enforced in developing countries. Despite this, recreational waters are widely used and leisure activities represent one of the main economic incomes for the local population. In tropical countries such as Ecuador, beaches are visited by tourists from all over the country and abroad throughout the year. The beaches are crowded especially on holidays, four of them (Christmas, New Year’s Eve, Carnival and Easter) occurring during the rainy season of the Ecuadorian Coast (December-April) making these coastal areas susceptible to runoff events. Ecuador has a total of 421 wastewater treatment plants; of these, 129 are located in the coastal region (INEC, 2016), however, the effectiveness of their treatments and capacity for the growing population is poorly known.

Ecuador has a total of 421 wastewater treatment plants, 129 of which are located in the coastal region (INEC, 2016). The majority of municipalities (62%) conduct wastewater treatment for their communities, while the remaining municipalities (38%) do not carry out any type of treatment (INEC, 2016). Playas canton has 41,935 inhabitants and Salinas 68,675 (INEC, 2010). The main economic activities in both cantons are fishing, and sun and beach tourism that take place throughout the year. These places receive national tourists in the months of December to April (coastal region holiday season); and the month from August to October (highland region holiday season). Santa Elena has a sanitary system that covers 60% of the urban area. This sanitary network collects wastewater to a pumping station and then towards oxidation lagoons, where they also receive influents from Salinas, Santa Rosa, Muey and La Libertad. Wastewater receives a secondary treatment with oxidation and subsequent disinfection with chlorine to be discharged into the sea while Playas has a facultative lagoon (Córdova, 2013).

The use of recreational waters promotes one of the main economic activities in Ecuador, therefore, the survey of its quality to guarantee users health should become a priority. The purpose of this study was to assess water quality and the presence of SARS-CoV-2 at the most visited beaches in Ecuador and at two oxidation lagoons at Guayas and Santa Elena during restrictions (during the dry season in 2020) and opening of beaches (during the rainy season in 2021).

## Materials and Methods

### Study sites

The study area comprised marine waters located between latitude 1° 56’ 5,81” S and 2° 43’ 24,368” S. and longitude 80° 43’ 24,6” W and 80° 18’ 18,493” W along the Ecuadorian coastal area. Fifteen beaches and two oxidation lagoons were selected along the Guayas and Santa Elena provinces. They included artisanal fishing ports, estuaries, recreational beaches and bays, and a diving area located 10 km from the coast. Description of the sampling sites is shown in Table 1. All areas were sampled during the Ecuadorian confinement established for reducing the impact of the COVID-19 pandemic during the dry season in July 2020 (A1-O2) and after the confinement in the rainy season in January 2021 (A12-O22).

**Table 1.**
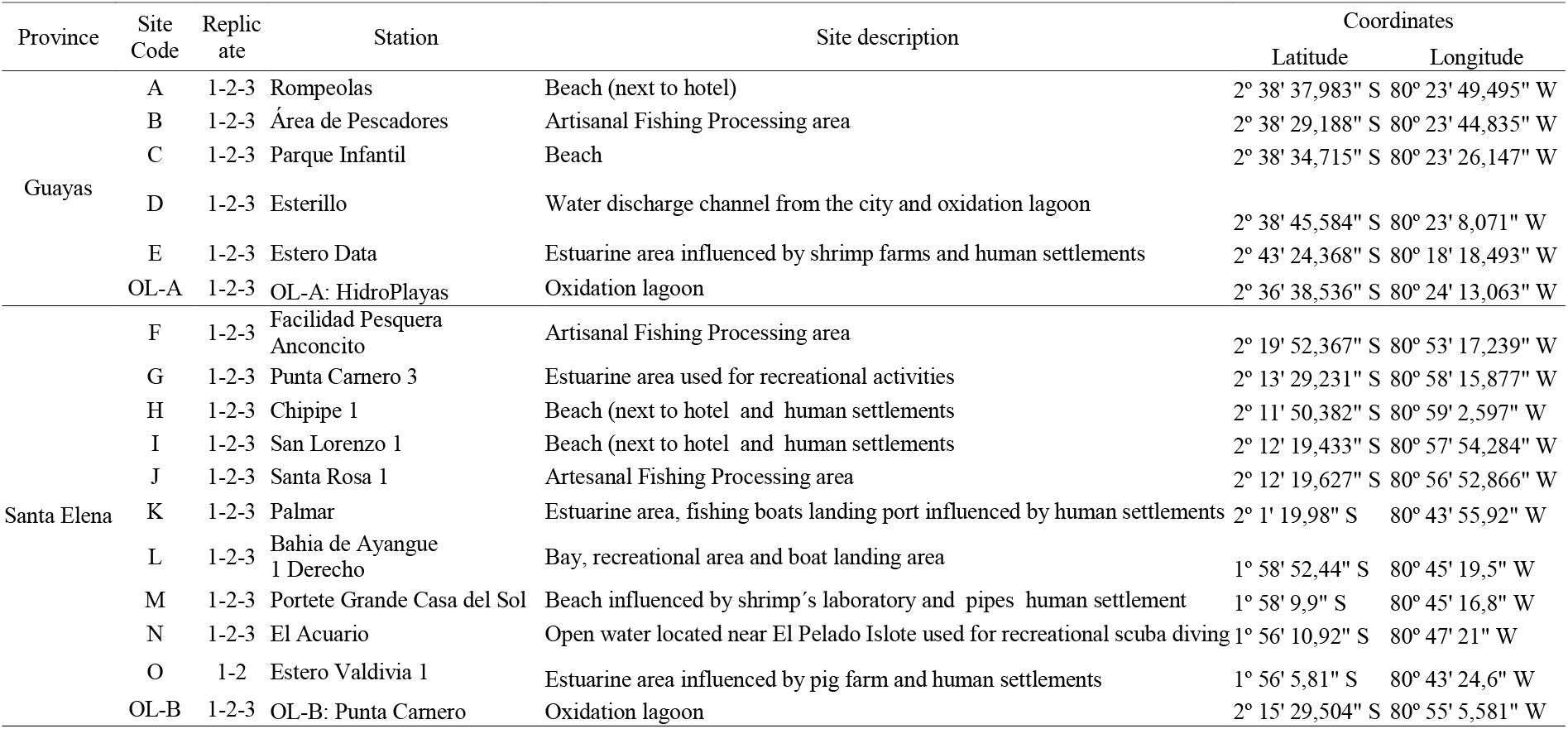
Sampling sites. Provinces, locations, site code, site description and coordinates from study sites.

### Water sampling and *in situ* analyses

A total of 84 surface seawater samples were collected at 15 beaches (A1-O22) and two oxidation lagoons, in Playas (OL-A) and Punta Carnero (OL-B) (Figure 1, Table 1) between July 18^th^, 2020 and January 30^th^, 2021. Water samples (1L) were collected during the low-tide. using a telescopic rod sampler at a depth of 30 cm using one liter sterilized plastic bottles. All samples were collected in triplicates, except for the Valdivia location due to logistical limitations. Samples were immediately put at 4 °C in the dark, and transported to the laboratory within 6-12 h of collection. For SARS-CoV-2 detection, the samples were stored at −80 C until further analysis at the Multidisciplinary Center for Research of the Ecuadorian National Health Institute (Ecuadorian NHI).

**Figure 1.**
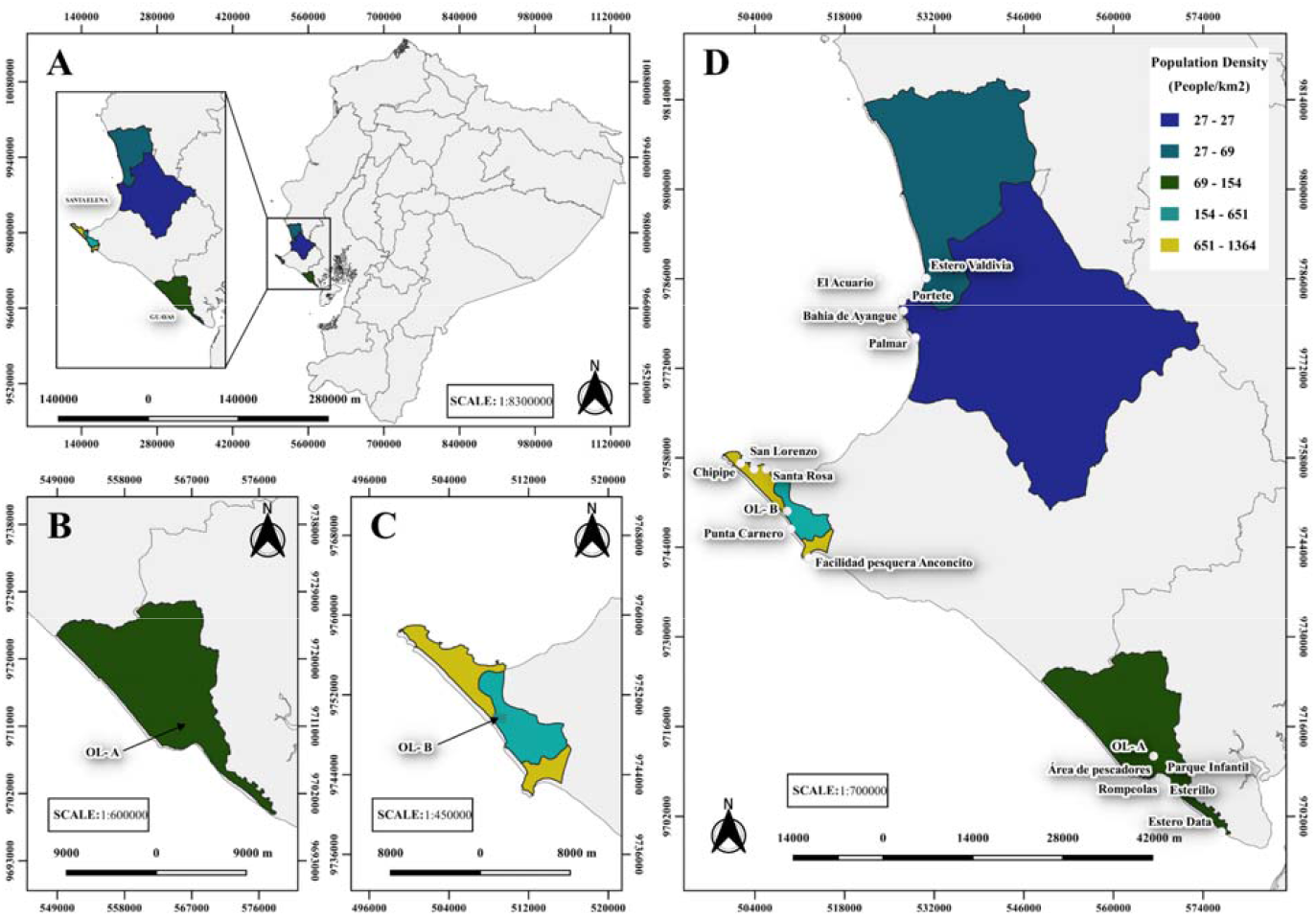
Sampling sites locations and population density of studied sites: (A) sample site location within Guayas and Santa Elena Provinces in Ecuador (B) oxidation lagoons of Playas (OL-A) situated in Guayas province (C) oxidation lagoons of Punta Carnero (OL-B) situated in Santa Elena province (D) the sites studied and its population density.

### Physico-chemical analyses

Throughout the whole sampling period, fourteen variables were measured. Of these, four were obtained *in situ* at each sampling site: Temperature (Temp), pH in water, salinity (Sal), and dissolved oxygen (DO), using YSI HQ30d multiparameter probes.

In the laboratory, seven parameters were measured: chemical oxygen demand (COD), biological oxygen demand (BOD), oils & fats (OF), surfactants or tensoactives (TENS), total residual chlorine (Cl), ammonium (NH4), total suspended solids (TSS), according to standard methods (Rice et al., 2017).

COD analysis was performed by VIS spectrophotometry at a wavelength of 620 nm using a HACH DR / 6000 spectrophotometer. The sample was previously digested for 2 hours at 150 ° C with sulfuric acid and a strong oxidizing agent (potassium dichromate). The BOD analysis was carried out by calculating the difference between the initial oxygen and the final oxygen determined in the sample after 5 days of incubation with a standard microbial community (Rice et al., 2017). The pH and Salinity were determined by electrometry using a glass electrode with temperature compensation, and a calibrated potentiometer, respectively. Surfactant analysis was performed by extraction in benzene from an acidic aqueous medium containing excess methylene blue, followed by countercurrent washing with water, and the determination of the blue color by spectrophotometry at 652 nm using the HACH DR / spectrophotometer 6000. The analysis of oils and fats was carried out by gravimetry by extraction with hexane (Rice et al., 2017).

TSS determination were performed by gravimetry after filtering the sample using a 0.45 um pore diameter filter, drying the residue on the filter and determining the weight gain. The analysis of residual free Chlorine was carried out by spectrophotometry using the HACH DR/6000 spectrophotometer, after the addition of the indicator N, N-diethyl-p-phenylenediamine (DPD). The ammonia analysis was carried out following Rice et al. (2017), a mineral stabilizer and the dispersing agent are applied to the sample that helps the formation of the reaction of the Nessler reagent with N-ammoniacal ions, forming the yellow color which is measured by spectrophotometry.

### Microbiological analysis

The microbiological parameters included *Fecal coliforms* (FC), *Escherichia coli* bacterial numbers (EC). The analysis of FC was performed using the multiple tube fermentation technique, estimating the bacterial density using the most probable number method (Rice et al., 2017). EC analysis was performed using the membrane filtration technique using the selective growth medium m-ColiBlue24 and incubating the samples at 44.5 ° C. Densities in both cases were determined by using Most Probable Number (MPN) method (Halkman, 2005).

### SARS-CoV-2 Analysis

#### Viral particle concentration and RNA isolation

The analysis of SAR-CoV-2 was conducted at two laboratories of the National Institute for Public Health Research of Ecuador (INSPI): 1) the Multidisciplinary Center for Research and 2) the Reference National Center for Exanthematic and Arthropod Borne Viruses. Water samples were processed into a biosafety cabinet Class II, Type A 2. One of the key factors for virus detection in water is the use of methods that allow the concentration of viral particles in enough amounts to be detected by the diagnostic test of interest. Therefore, two different concentration methods were evaluated: 1) skimmed milk organic flocculation (SMF), which is based on the capacity of the proteins to form flocs where the viruses present in the samples usually adhere (Calgua et al., 2008) and 2) concentration of organic particles with an aluminum chloride solution (AlClL), based on viral adsorption– precipitation (Randazzo et al., 2019). A brief modification was introduced for the SMF method: 1000 mL was used as initial sample volume instead of 10 L, and ∼ 100 mL of water containing flocks were processed for the centrifugation steps. To test whether the concentration methods were effective in concentrating enveloped viruses, a sterile water sample was spiked with Dengue Virus serotype 2-(DENV 2) isolated from C36/ cells. A 1:10 serial dilution set of this spiking control was evaluated for SMF (1:10 up to 1:1000), and a sole 1:100 dilution was used for AlClL. All supernatants from the different stages of the viral concentration procedure were treated with chlorine overnight prior to its elimination through the laboratory’s piping. Water samples were also evaluated without pretreatment for concentration of viral particles, in this case 1000 mL of sample was used for this purpose. RNA was extracted using commercial QIAamp Viral RNA Mini Kit (Qiagen Sciences, Germantown, MD, USA) following the manufacturer’s instructions.

#### Quantitative reverse transcription PCR (RT-qPCR) analysis

Analysis of water samples spiked with DENV 2 was carried out using a one-step PCR procedure. For DENV, the commercial Simplexa™ Dengue Kit was used (FOCUS Diagnostics, Cypress, CA, USA). A monoplex reaction for detecting DENV 2 was conducted, reactions were run in a 3M Integrated Cycler real-time RT-PCR instrument (3M-FOCUS Diagnostics). Survey of SARS-CoV-2 RNA was performed using the CDC 2019-Novel Coronavirus (2019-nCoV) purchased from Integrated DNA Technologies, Inc. (Coralville, IA, USA). The RT-PCR assay was set up as described by the manufacturer using a SuperScript III Platinum One-Step qRT-PCR kit (Invitrogen, Waltham, MA, USA). The CDC protocol (CDC, 2019) targets two genes of the viral nucleocapsid (N1 and N2), the kit includes a positive control for SARS-CoV-2 and for human ribonuclease P (RNase P). Reactions were run on a BIO-RAD CFX96 Real-Time PCR System, with the following thermocycling conditions: one cycle of 50 °C for 30 seconds, one cycle if 95 °C for 2 minutes, and 45 cycles of 95 °C for 15 seconds and 60 °C for 1 minute. As for the CDC protocol, a RT-PCR amplification with CT <40 for both genes N1 and N2 was considered positive, amplification of only one gene was considered inconclusive and no amplification was considered negative. Amplification of RNase P gene is not expected from environmental samples. RNA samples leading to positive results were sent for confirmation to the National Reference Center for Influenza and other Respiratory Viruses of the INSPI, a laboratory fully accredited and certified by the World Health Organization for diagnosis of SARS-CoV-2. This laboratory uses the protocol of the University of Charité (Corman et al., 2020). Analysis of samples was carried out targeting a gene of the viral envelope (E).

### Statistical analysis

The data exploration was done using draftsman plot. Kolmogorov-Smirnov test was conducted for the analysis of normality of data distribution, and the Levene test for homoscedasticity (Zar, 1996). Subsequently, given the lack of normality for the whole data set, a non-parametric test as Kruskal-Wallis was performed (Sokal and Rohlf, 1995) to identify statistical differences between sampling sites regarding water physicochemical and microbiological parameters.

The entire sampling seasons were considered to calculate maximum, minimum and mean values of physical and chemical parameters. Multivariate analyses were performed using the statistical package Primer-E (Clarke and Gorley, 2006), version 7.0.20. Multivariate analysis of all sampled sites was carried out based on physicochemical and microbiological parameters (Clarke and Gorley, 2006) only for the data collected in beaches. The Euclidean distance was used to construct a similarity matrix from the fourth-root transformed data for each replicate sample at each site in each site and season. The matrix was then subjected to Non-Metric Multidimensional Scaling (n-MDS) ordination previous individual transformation Log (V+0.1) of the variables FC, EC, TENS and TSS. Two-way crossed ANOSIM tests were used to identify significant differences between season and sites. In each test, the null hypothesis that no significant differences are found between season and sites was rejected if the significance level was p≤0.001. The R statistic value was used to ascertain the extent of any significant differences (Clarke and Warwick, 2001). Similarity percentages test (SIMPER) was used to identify which variables made the greatest contributions to those differences. Finally, the distribution of FC in the coastal areas and OL and pollutant critical points were plotted on the Ecuadorian map using QGIS program version 3.4.3.

Results of water quality were compared with the current Ecuadorian environmental regulations by use of the Book VI from the Unified Text of Secondary Legislation of the Ministry of the Environment (TULSMA): Environmental Quality Standard and effluent discharge to the Water Resource (Acuerdo Ministerial 097A) for *Admissible quality criteria for the preservation of aquatic and wildlife in fresh, marine and estuarine waters* (Table 2); *Water quality criteria for recreational purposes through primary contact* (Table 6) and *Limits of discharges to a body of seawater* (Table 10).

**Table 2.**
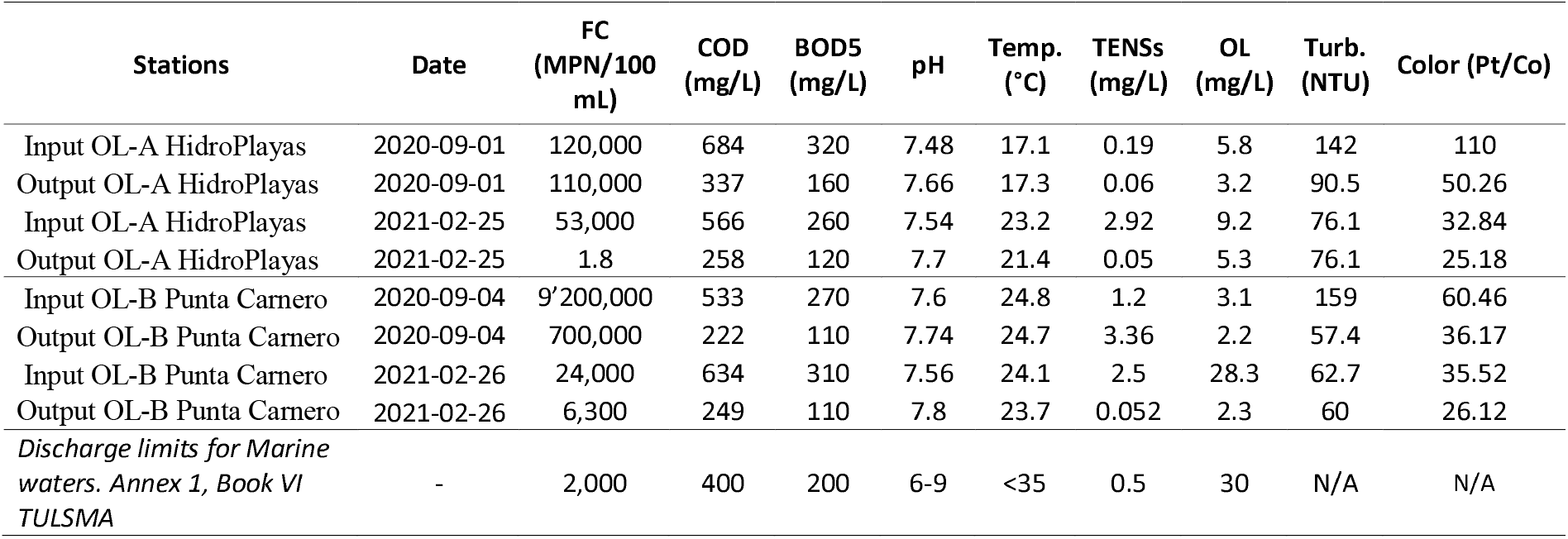
Microbiological and physicochemical parameters of oxidation lagoons at Guayas and Santa Elena.

## Results

### Water physicochemical parameters

Maximum, minimum and mean values obtained from physical and chemical parameters at each sampling site for dry season (2020) and rainy season (2021) are shown in Table S1. The temperature ranged from 23.6 °C to 34.4 °C with higher values in the rainy season (January) and lowest in dry season (July), due to differences between the season period and time of survey (early in the morning or past midday). A higher temperature was observed in Estero Valdivia and Esterillo. The higher TSS were observed at Ayangue in the Estero Valdivia (698 mg/L) in an area close to a pig farm, Fisherman area (114 mg/L) and Esterillo (86 mg/L) located near restaurants in Playas, mainly in the rainy season (Table S1). Dissolved oxygen concentration fluctuated between 2 to 9 mg/L, with higher concentration in the rainy season (July) and lowest concentration associated with the estuarine area called Estero Valdivia and Area de Pescadores. Concentrations lower than 5 mg/L (below the recommended concentration for the preservation of the flora and fauna) were found at fishing facilities (Anconcito), estuaries (Esterillo and Valdivia) and fishing ports (Area de Pescadores). COD ranged from 27mg/L to 820 mg/L. The highest values were registered at the outer and middle branches of Estero Valdivia during the rainy season; these values exceeded the permissible COD limits for the preservation of flora and fauna according to environmental laws in Ecuador (400 mg/L). Other sites with high levels although within limits were found at the estuarine zones such as: Estero Data 1 (384 mg/L), Estero Data 3 (302 mg/L), Palmar 3 (260 mg/L) and Punta Carnero (210 mg/L) (Figure 2). BOD ranged from 2 to 200 mg/L, higher levels were observed only at Estero Valdivia in the middle (150 mg/L) and outer estuary (200 mg/L) during the rainy season (Figure 2).

**Figure 2.**
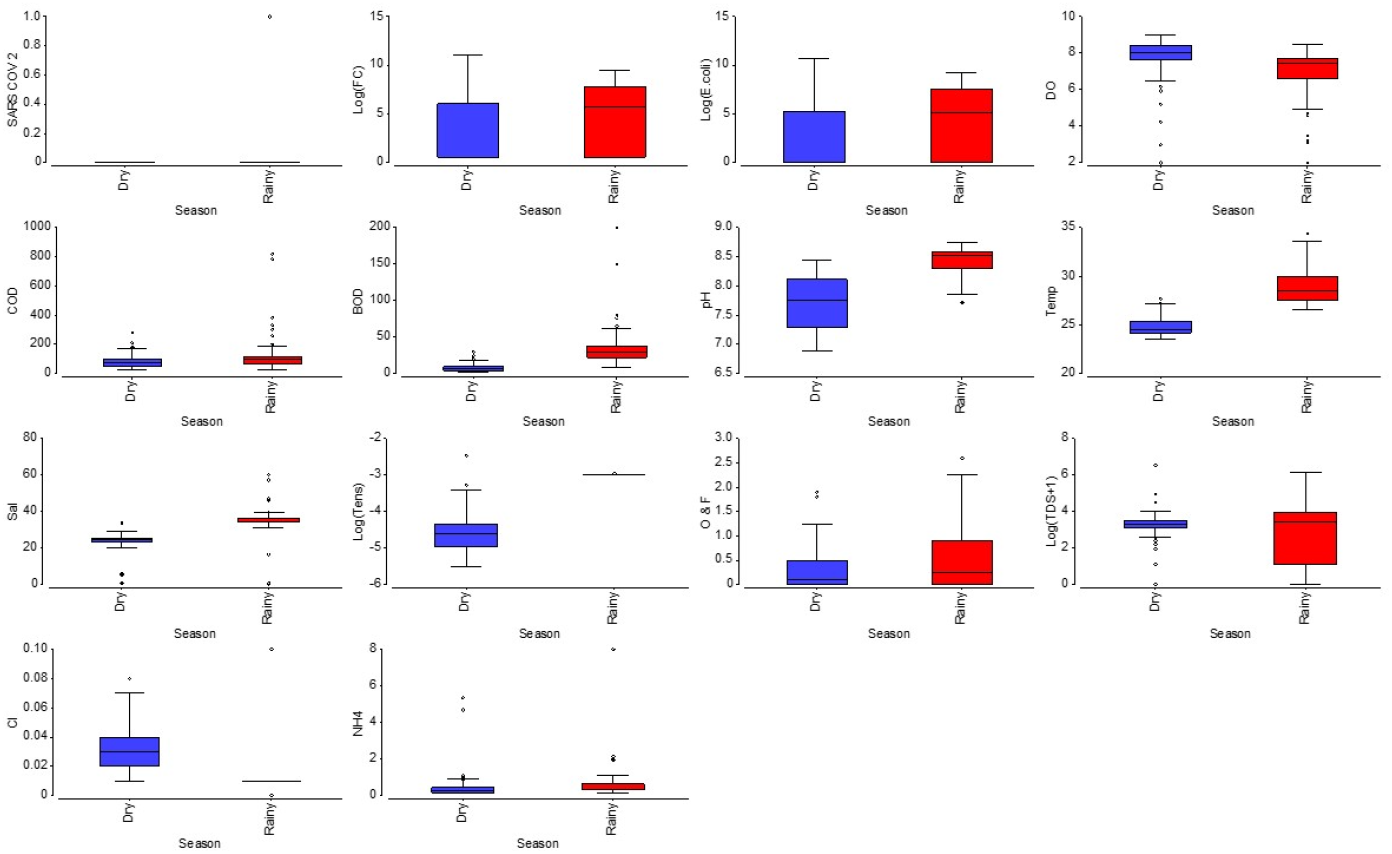
Physico-chemical and biological quality parameters assessed in seawater of beache during dry (blue) and rainy (red) seasons. FC: fecal coliforms; DO (mg/L): dissolved oxygen; COD (mg/L): chemical oxygen demand; BOD (mg/L): biological oxygen demand; temp (Celsius): temperature; Sal (Practical salinity units): salinity; tens mg/L): tensoactives; O & F (mg/L): Oils and fats; TDS (mg/L): total dissolved solids; Cl: Chlorine (mg/L); NH_4_ (mg/L): ammonium.

The pH ranged from 6.88 to 8.75, the lowest values were in the July of 2020 at Estero Data (6.88) and in the rain season (7.72) at Esterillo. Salinity ranged from 0.5 to 60.2 PSU; an alteration in salinity was observed in intertidal zones due the influence of wastewater from Playas county, and in landing and commercialization of fishing in Area de Pescadores; estuarine area (Esterillo). The highest salinity values were found in the external area from Estero Valdivia where high temperatures increase evaporation rate, increasing salinity in the area near the mouth of the sea during the rainy season. Tensoactives showed values between 0.004-0.085 mg/L, the highest concentrations were observed in Punta Carnero 1 during February 2021 (Table S1). In addition, ammonium ranged from 0.12 to 8 mg/L with highest levels at intertidal zones where domestic wastewater is discharged (Area de Pescadores), and estuarine zones such as Esterillo and Estero Valdivia (Table S1). The concentrations of residual free chlorine and surfactants were less than the detection limit <10 mg/L and <0.05 mg/L, respectively. There were only significant (p <0.05) differences between sampling sites for Temp, Sal and OF. In this context, OF ranged from 0.07 to 2.6 mg/L, exhibiting the highest values in rainy season (January 2021). OF values tended to be higher (≥ 2 mg/L) in fishing boats and landing ports where the artisanal fishing fleet is sheltered (Anconcito, Portete Grande, Area de Pescadores and Estero). This increase coincided with the beaches that were reopened with higher tourist activity. Similar trends were observed in water channels with wastewater and industrial runoff coming from Playas’ downtown discharged in the intertidal zone at Esterillo.

A two-way crossed ANOSIM demonstrated that water quality was influenced by both site and season (R= −0.76; *p=0*.*001*).

### Microbiological assessment

FC counts were very variable, ranging from 1.8 to 63,000 MPN/100 mL. The highest levels were found in samples collected at artisanal fishing ports, estuaries, and urbanizations. Although the highest value was found during the dry season at Anconcito Artisanal Fishing Port, in general the higher levels and number of sites were observed in the rainy season (Figure 3).

**Figure 3.**
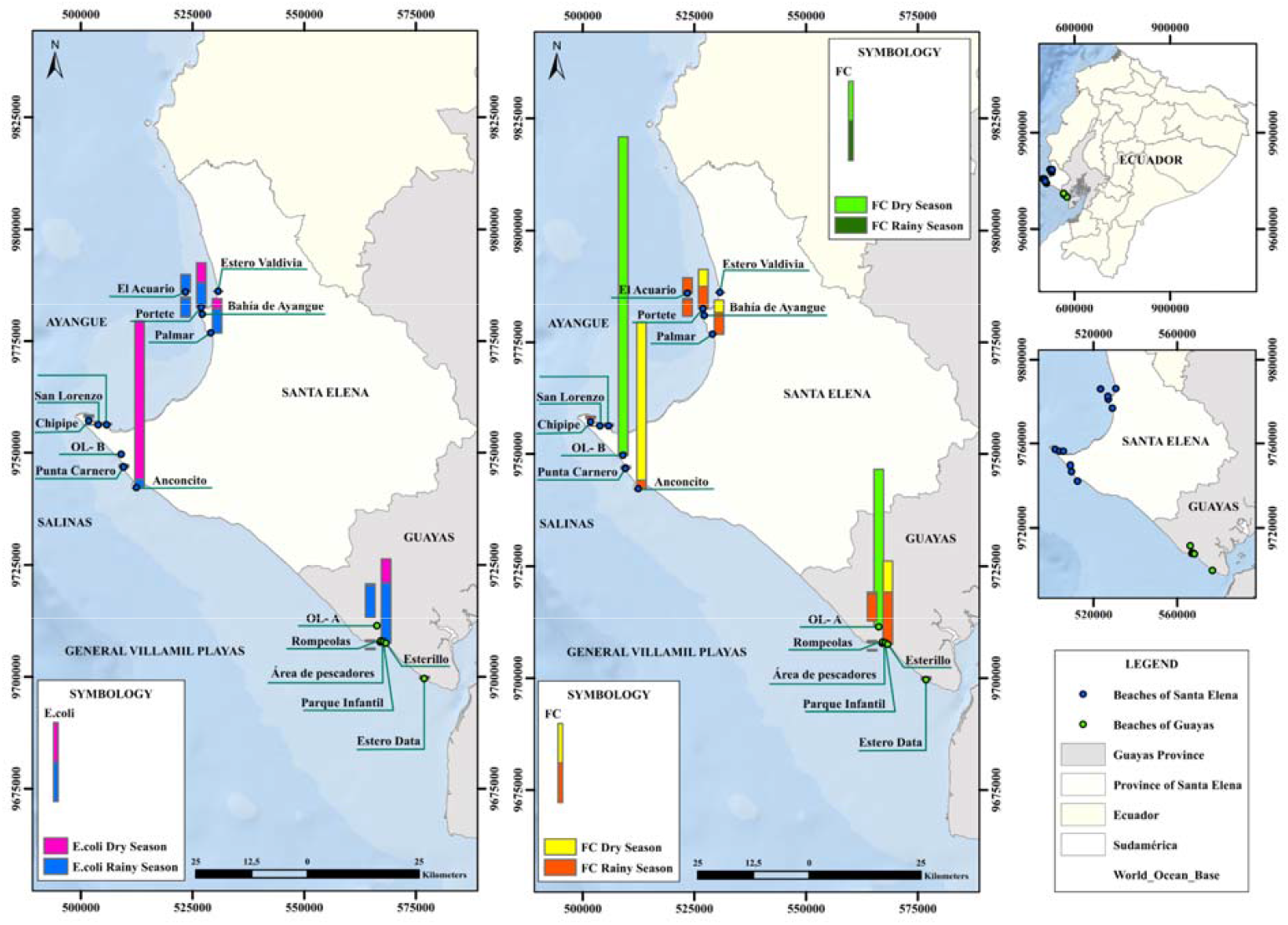
Spatial distribution of *E. coli* (left) and fecal coliforms (right) in the seawater and wastewater treatment systems during 2020-2021. The concentration of fecal coliforms in OL-A and OL-B are light green in Dry season and dark green in the rainy season. Fecal coliforms on beaches are yellow in the dry season and orange in the rainy season.

Both FC and EC were consistently higher in seawater and wastewater discharged in estuarine zones compared to shoreline sand. Values were high at a fish landing and management facility located at Anconcito Artesanal Fishing Port, at a channel of domestic wastewater (Esterillo 1 and 2), in areas next to restaurants and public toilets, on the coastline near to urbanizations (Portete Grande Casa del Sol), and an estuarine area influenced by urban settlement (Palmar 1) (Figure 3). The highest FC values were detected in Anconcito (63,000 MPN/100 mL), Esterillo (9,200 MPN/100 mL), Portete (7,000 MPN/100 mL) and Palmar (4,000 MPN/100 mL). The same trends were registered for EC (Figure 3).

There was a statistically significant difference between sites for FC (p=0.03) and EC (p=0.03). Also, both FC and EC were higher (p<0.05 and p=0.01, respectively) in the rainy season when beaches opened for tourism compared to the dry season and closed beaches. FC were highest in Santa Elena in relation with Playas (p<0.05 and p=0.01) in both survey times. However, a higher number of sites showed water polluted with FC and *E coli* in the area from Ayangue, which is a preferred area by tourists due to its calm waters especially in Ayangue Bay and surroundings. In fact, only ten of the surveyed sites could be suitable for recreational purposes according to environmental laws in Ecuador through ministerial agreement 097A (Acuerdo Ministerial 097A, 2015). Here, it is recommended that for recreational purposes, waters should not exceed 200 MPN/100 mL. These sites were Rompeolas, Parque Infantil 2 and Estero Data 1 in Playas; Punta Carnero 1 and 2, Chipipe 1 and 2; San Lorenzo 1,2,3 and Estero Valdivia in Santa Elena.

### Relation between seawater water quality and environmental factors

The non-metric multidimensional scaling ordination showed that there was differentiation of the water quality generated by seasonality between the dry and rainy seasons (Figure 4). The similarity of studied sites was influenced by the levels of TENS, Temp, pH, BOD, Sal, Cl, EC, an DO. The SIMPER routine showed the variables with most contribution were TENS (10.54%); Temp (9.94%); pH (8.87%), BOD (7.46%) and salinity (7.15%).

**Figure 4.**
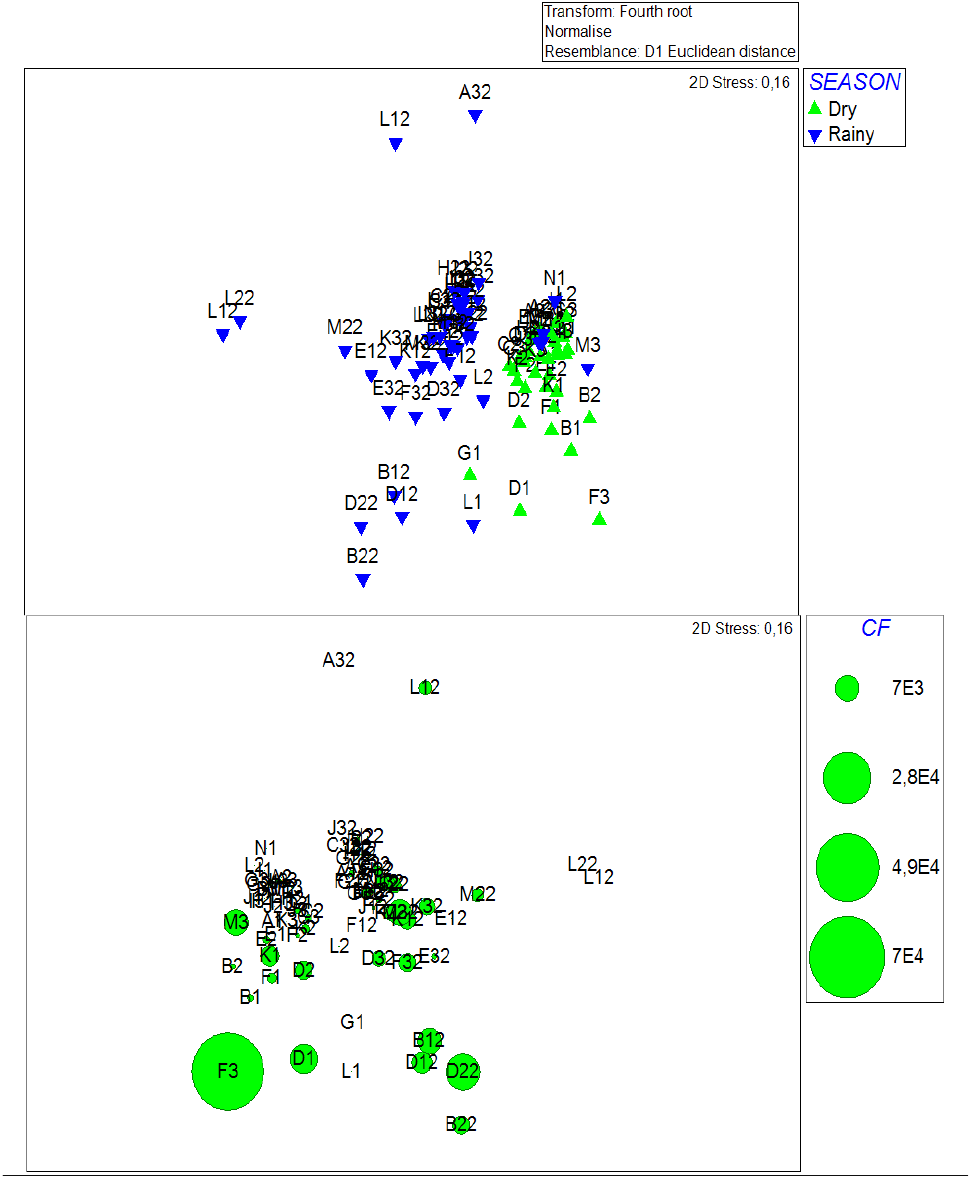
Non–metric multidimensional scaling ordination, derived from Euclidean distance constructed using square root transformed total chemical, physical and microbiological parameters at 42 sampling sites during dry season (2020) and rainy season (2021). The upper plot has used for all variables and seasons and is coded for season, while the lower plot were restricted to Fecal Coliforms for all seasons.

### Microbiological pollution in Wastewater in Oxidation Lagoons

The main microbiological, physical and chemical parameters in OLs are shown in Table 2. The concentration of FC in the oxidation lagoons fluctuated between 1.8 to 9’200,000 MPN/100ml. The highest levels were showed in Santa Elena during 2020, with maximum levels registered in the entrance of Punta Carnero (OL-B, 9’200,000 MPN/100 mL) while lower concentrations were recorded in the Playas’s lagoons (120,000 MPN/100 mL). The levels observed in the output of both oxidation lagoons were high, with values above the allowed limit (2,000 MPN/100 mL) for discharge from wastewaters to seawater in Ecuador. The high levels of FC recorded at the output of the oxidation ponds analyzed show low performance and low efficiency in the removal of organic matter and microorganisms in these systems.

### SARS-CoV-2 Analysis

The two viral concentration methods evaluated, SMF and AlCl_3,_ led to the detection of DENV-2 in all dilutions tested, suggesting both procedures are effective to adhere enveloped viruses such as coronaviruses. Due to the shorter processing time, the AlCl_3_ procedure was selected for the analysis of SARS-CoV-2 in the water samples. SARS-CoV-2 RNA was surveyed on samples from two oxidation lagoons in Playas (OL-A) and Punta Carnero (OL-B) and five coastal waters exceeding permissible limits of FC. The results of the RT-PCR are shown on Table 3. In samples collected during confinement, genes N1 and N2 of SARS-CoV-2 were detected in influent and effluent samples from both oxidation lagoons. Findings were confirmed by the National Reference Centre for Influenza and other Respiratory Viruses through amplification of the viral gene E, a laboratory certified by the World Health Organization for SARS-CoV-2 diagnosis. Gene N1 was amplified from RNA extracts collected at two artisanal fishing ports (Area de Pescadores, Facilidad Pesquera Anconcito) and one estuarine area (Palmar). After confinement, N1 and N2 genes were detected at one recreational beach (Bahía de Ayangue), only one gene was amplified from samples of one recreational bay (Portete Casa Grande del Sol), and both genes in both of the oxidation lagoons sampled. In accordance with the CDC guidelines, the results for the samples where only one gene was amplified was considered inconclusive. Of the samples with inconclusive results, two samples were also processed using the SMF method. RT-PCR confirmed the results as inconclusive. As expected, the RNase P gene control did not yield any amplification.

## Discussion

Water quality parameters revealed a negative impact on the beaches studied associated with human activities. Fecal coliforms and *E. coli* values exceeded the maximum permissible levels of Environmental Quality Standard and effluent discharge for water sources (Ministerial Agreement 097A). Water quality criteria for recreational purposes through primary contact, and preservation of aquatic and wildlife in fresh, marine and estuarine waters in areas influenced by fishing ports, wastewater, urbanizations were also exceeded [Annex 1 of Book VI from Unified Text of Secondary Legislation of the Ministry of the Environment (TULSMA) (Acuerdo Ministerial 097A, 2015)]. Moreover, SARS-CoV-2 RNA was found in oxidation lagoons included in this study and two natural water bodies.

Notably, the majority (9 out of the 15) of the beaches analyzed were not suitable for primary contact and/or recreational activities, due to biological contamination with fecal coliforms and *E. coli*. This pollution may have been generated by discharges of domestic wastewater, runoff, contaminated water from rain or subsoil by seepage, river mouths or direct bather shedding into seawater (Badilla-Aguilar et al., 2019). It is important to mention that this research was conducted during COVID-19 pandemic, before a vaccine was available, and while the use of beaches was prohibited. Therefore, it is possible to infer that microbiological contamination could be even be greater during holidays when there is a greater influx of tourists. The fact that so many beaches studied were contaminated with *E. coli*, fecal bacteria associated with gastrointestinal disease, represents a risk to human health due to the possible presence of other microorganisms including ESKAPE pathogens, protozoa and enteric viruses in these aquatic ecosystems (Li, et al., 2021; Nishiyama et al., 2021; Omarova et al.,2018; Rodrigues et al., 2016). Anconcito and Esterillo were the beaches with the highest pollution in terms of fecal coliforms. A possible explanation for these highly contaminated beaches may be associated with higher significant fecal coliform counts and high chemical oxygen demand observed in the raw sewage and oxidation ponds of these locations.

This study revealed that the amounts of coliforms were higher at the inlet and outlet of oxidation ponds during the beach closure season compared to the beach opening season when confinement measures were lifted. The oxidation ponds are highly efficient when operating under efficient conditions of residence time and organic load. However, the coliform data in the studied oxidation ponds suggest that an evaluation and improvement of the methods used to eliminate bacteria and viruses in these systems is required. Even though the levels of coliforms were very high in the oxidation ponds, they were lower on the beaches where these waters were eventually discharged, and the presence of the SARS-Cov-2 virus was not detectable, probably due to the dilution effect in the sea and to salinity conditions, incidence of ultraviolet radiation and the temperature of seawater (Liu et al., 2020). In this sense, it has been described that the survival of microorganisms in the environment depends on many factors such as temperature, incidence of sunlight and the presence of native microorganisms (Pinon and Vialette, 2018).

Another impact of the discharge of inefficiently treated wastewater to the beaches was the low levels of oxygen found (less than 5 mg/L). Due to the organic load of wastewater, effluent discharged from wastewater treatment facilities often contribute to the level of oxygen demand of the receiving water, due to the greater depletion of dissolved oxygen in the surface waters that receive water with poorly treated residuals (Huang et al., 2020). For example, DO levels in effluents from various wastewater treatment facilities in South Africa were typically lower than the required standard of 8 to 10 mg/L, and this condition was shown to negatively affect the aquatic ecosystem (Edokpayi et al., 2017). High levels of COD were observed in the beaches near urbanizations, fishing facilities, estuaries and bays, whilst highest levels of BOD were found in the Estero Valdivia showing there is organic contamination in water, which can also contribute to lowering DO levels due to bacterial respiration (Devi and Dahiya, 2008). The effect of poorly treated wastewater on surface waters is largely determined by the oxygen balance of the aquatic ecosystem, which is essential to maintain biological life within the system (Edokpayi et al., 2017). Therefore, there could be negative impacts of the lagoon systems in Playas and Santa Elena, impacting the local flora and fauna of these sites, which coincides with previous studies carried out in Playas (Espinoza, 2016) and Santa Elena (Córdova, 2013).

Finding of SARS-COV-2 RNA at the inlet of the oxidation lagoons sampled was expected as the virus is excreted in feces, but positive qPCR results in the effluent suggest that its treatment is not effective to eliminate the virus. Viral RNA was also found in two natural sampling sites, confirming fecal contamination is reaching the natural ecosystem. Nevertheless, RNA detection does not imply virus viability, for SARS-CoV-2 persistence of infectious particles in water matrices is temperature dependent (Sala-Comorera et al., 2021). In tropical countries it could also be affected by other environmental variables including seasonal seawater temperature, exposure to UV radiation, salinity, among others. The rapid decay of SARS-CoV-2 in environmental water supports the suggestion of the CDC that viral transmission through contact with contaminated waters is low. This result, however, highlights the possibility that other fecal derived pathogens with longer persistence rates could be introduced to recreational water. These might include gastrointestinal viruses known to be transmitted by the orofecal route such as noroviruses, and antibiotic resistant microorganisms which could pose a risk of transmission to users of these waters and to public health (Seitz et al., 2010, WHO 2021).

High levels of salinity registered in the Estero Valdivia can be attributed to the fact that there is not enough exchange in the Estero, and evaporation causes the salinity to increase, especially during low tide. The low levels of salinity generated by the wastewater that is discharged into the intertidal zone in the Playas area are probably related to clandestine connections that throughout the year are affecting biological communities, especially benthic assemblages that inhabit the estuarine area of the Esterillo and sandy beaches in the area where fishermen land (Scott et al., 2019).

Developing countries such as Ecuador have weak regulations for water quality, and surveillance of recreational water is not in place. The use of recreational waters promotes one of the main economic activities in Ecuador, survey of its quality to guarantee users and fauna health should become a priority. However, Ecuador as well as other countries in South America discharge untreated sewage polluting recreational use beaches, agricultural and fishing products that are grown in nearby areas, and negatively impacting economic activities as well as increasing risks to consumers’ health. It has been estimated that diarrhea accounts for one in eight deaths among children younger than 5 years per annum in South America, Africa and Asia (Kotloff, 2017). Diarrhea are due to viral, bacterial and protozoan species (Platts-Mills et al., 2015; Ugboko et al., 2020). Rotaviruses and *E. coli* are the most reported enteropathogens in the world, mainly in low-income countries (Onanuga et al., 2014). These organisms are associated with contaminated water and food. Our results emphasize the need for secondary or biological treatment such as activated sludge or moving bed bioreactors, rather than primary/physical treatment as conducted in these areas with oxidation ponds. Moreover, there is a need to develop an education program for owners of hotels and restaurants at tourist beaches, regarding good practices in food manufacturing and prevention of food-borne diseases (ETA).

## Conclusions

In this work, water quality of seawater and wastewater discharges in two regions of Ecuador were analyzed during two seasons during the COVID pandemic. It was found that 81% of beaches were not suitable for recreation, as they exceeding the maximum limit allowed in water for recreational activities, according to Ecuadorian environmental legislation. The virus SARS-CoV-2 was present in seawater and outfalls of estuaries, highlighting the need to reinforce prevention actions that guarantee the health of the users of these ecosystems. Fecal contamination could lead to the introduction of other human enteric viruses into aquatic ecosystems. Therefore, it is recommended to implement a water quality monitoring program including physical, chemical, and microbiological parameters (including the presence of SARS-CoV-2), especially in wastewater to avoid possible bacterial or viral outbreaks to residents and tourists.

## Supporting information

Supplementary Table S1: Physicochemical parameters in seawater of the beaches under study.

## Data Availability

All data produced in the present study are available upon reasonable request to the authors.

## Acknowledgements

The authors thank authorities of Prefectura de Santa Elena for the support with the fundraising for the sampling development in Santa Elena in 2020. Supplemental support was provided by the Secretary of Higher Education Science, Technology and Innovation from Ecuador (SENESCYT) and La Deutsche Gesellschaft für Internationale Zusammenarbeit (GIZ) DC program Sustainable Intermediate Cities in Ecuador. Component 3 “Strengthening of the Academy and Applied Research Institutes’’ grant No. 14.2160.1-001.00. We thank Ely Soares and Philipo Franco from the Municipality of Playas for provision of logistics and transportation of researchers to Playas in time of a COVID-19 pandemic. Special thanks to authorities from Ministerio del Ambiente, Agua y Transición Ecológica for the authorization to carry out research in marine protected areas. To Fanny Condo and María Belén Delgado from the Ministry of Tourism to cover costs of the maritime transportation at the Reserva Marina El Pelado. We also thank Doménica De Mora and Maritza Olmedo from the National Reference Centre for Influenza and other Respiratory Viruses confirming SARS-CoV-2 results. Finally, a posthumous recognition to Carlos Muñoz (+), Dean of the Faculty of Chemical Engineering of the University of Guayaquil, who unconditionally supported all the activities to conduct this study. A Visual and Automated Disease Analytics (VADA) Graduate Fellowship was awarded to Jhannelle Francis (University of Manitoba).

## Supplementary material

**Table S1.**
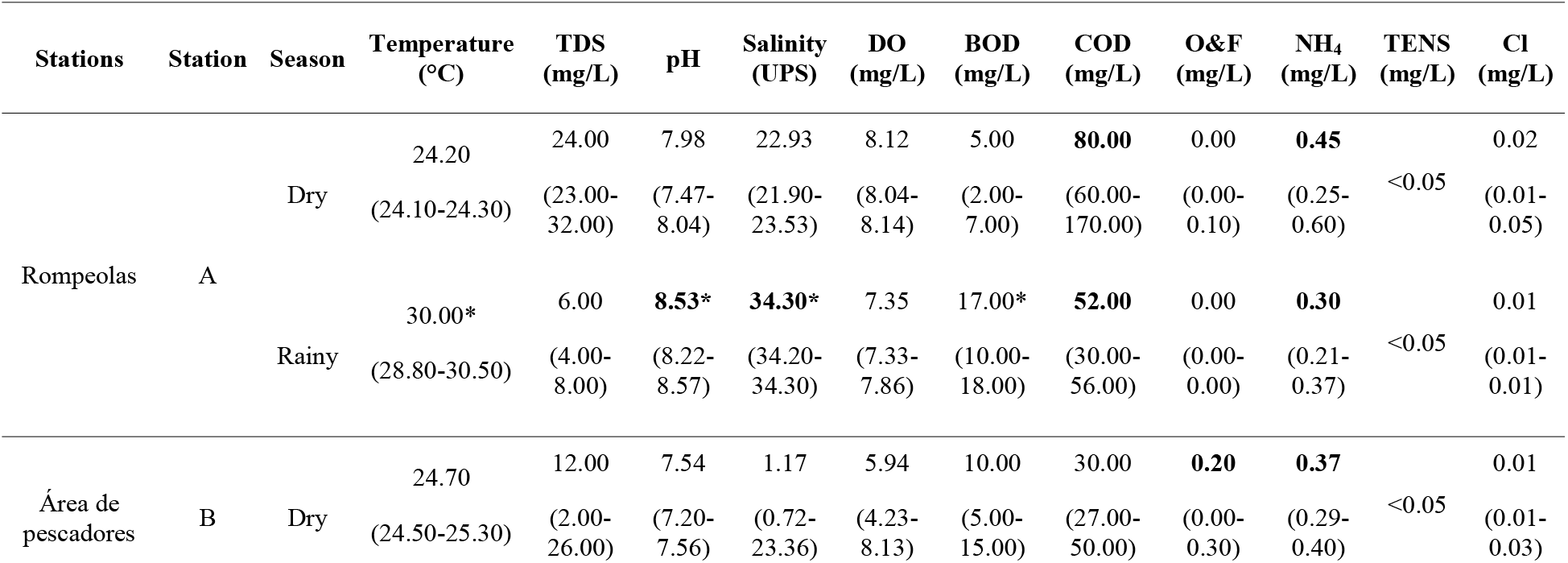
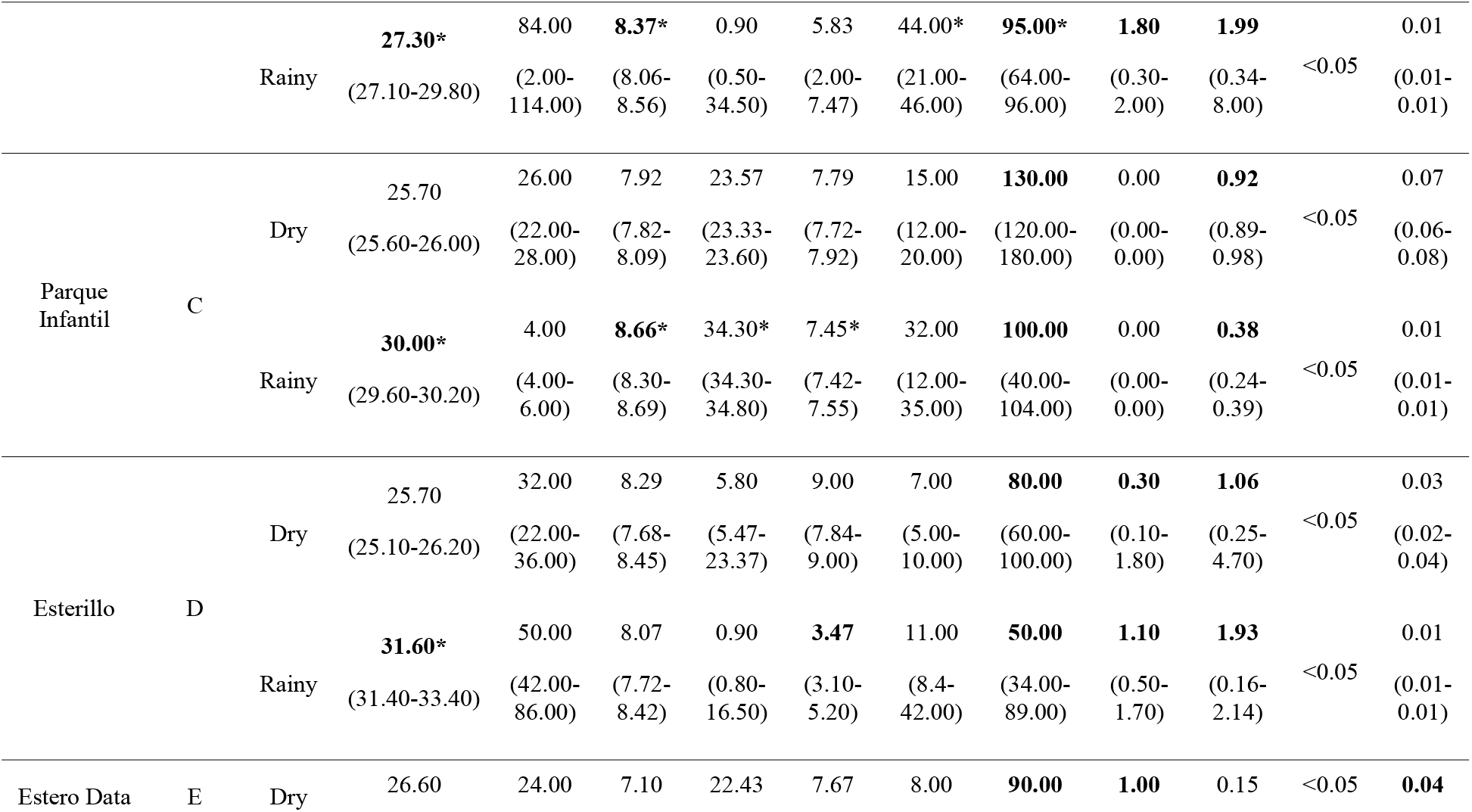
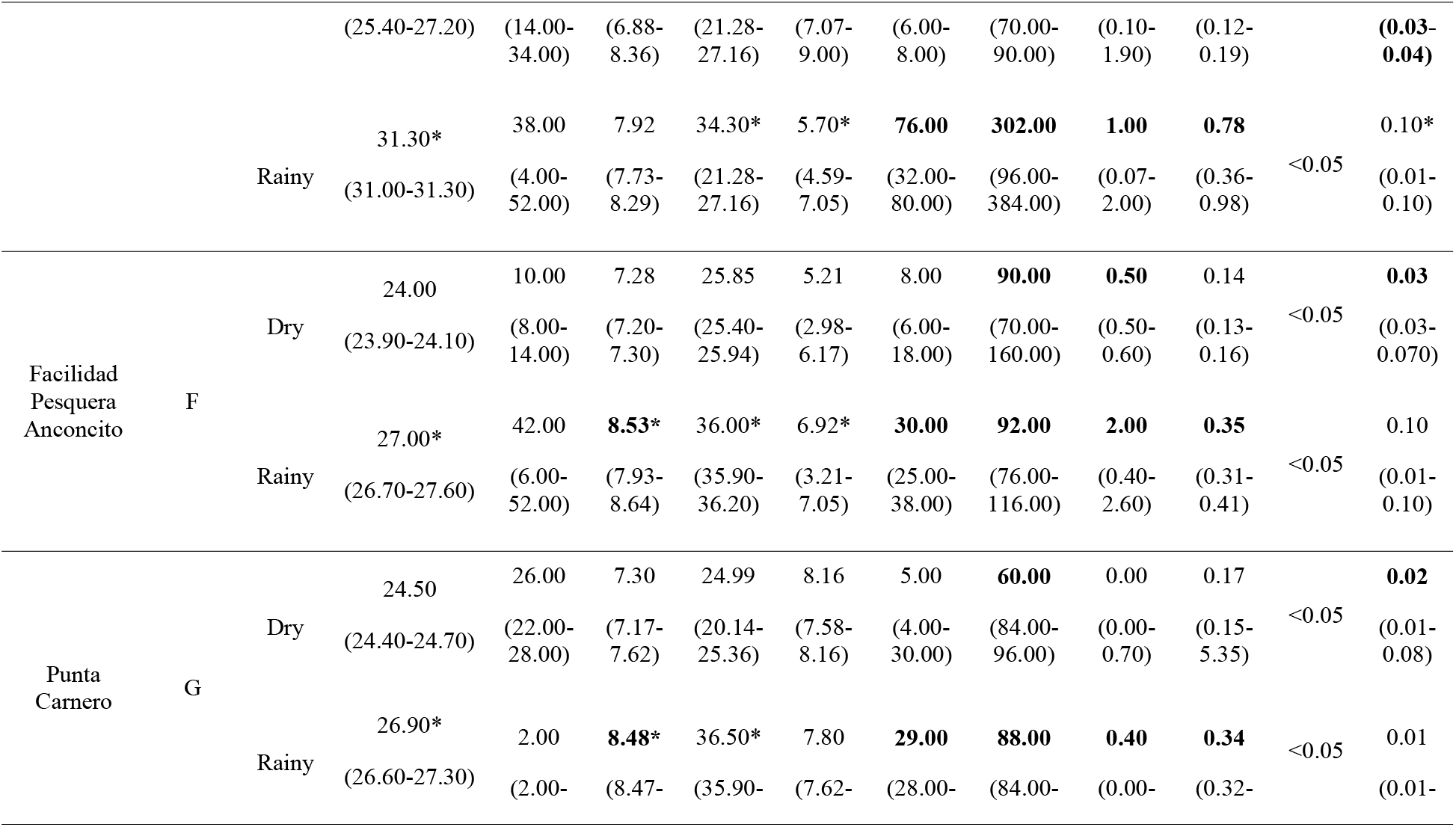
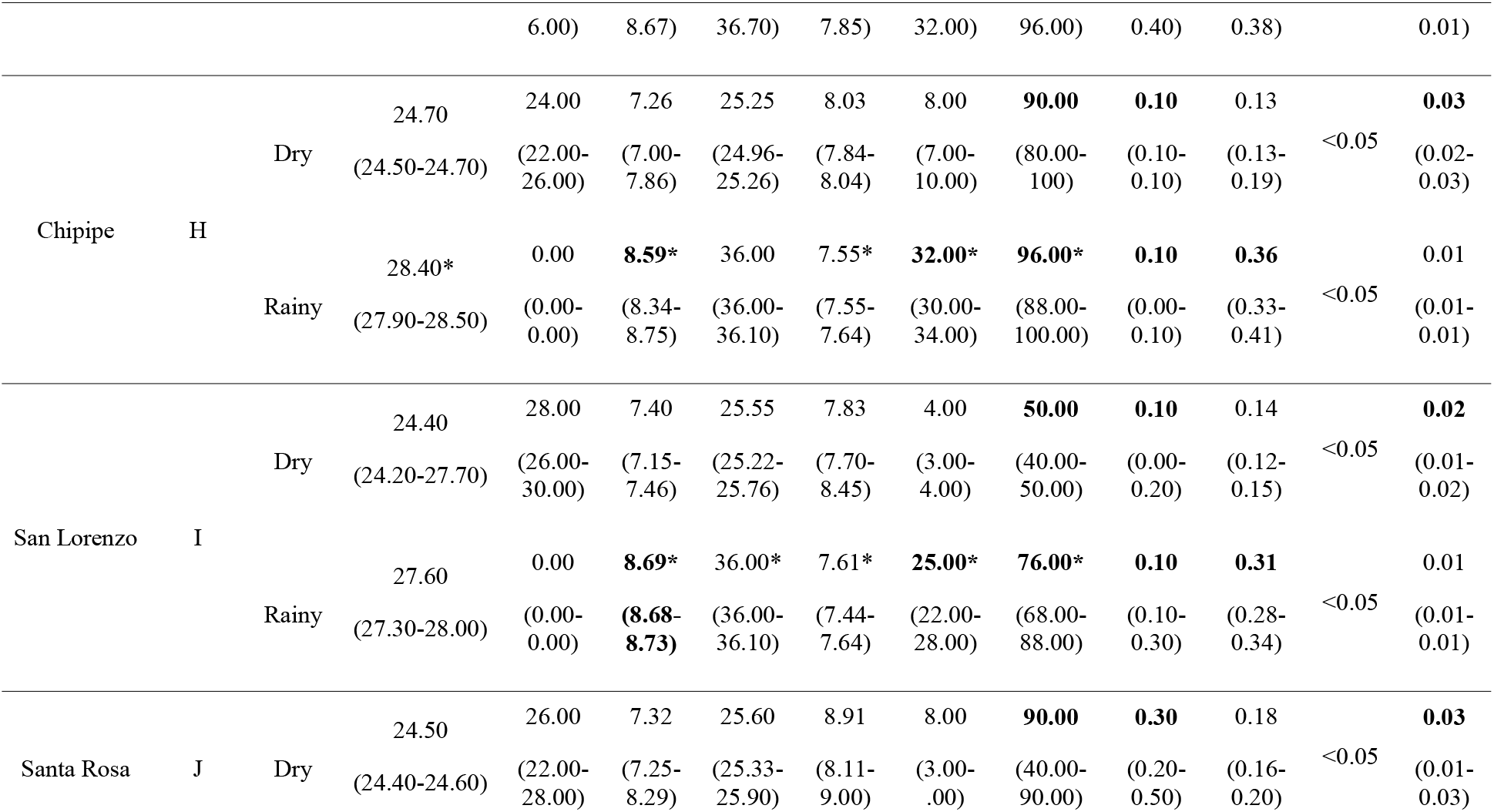
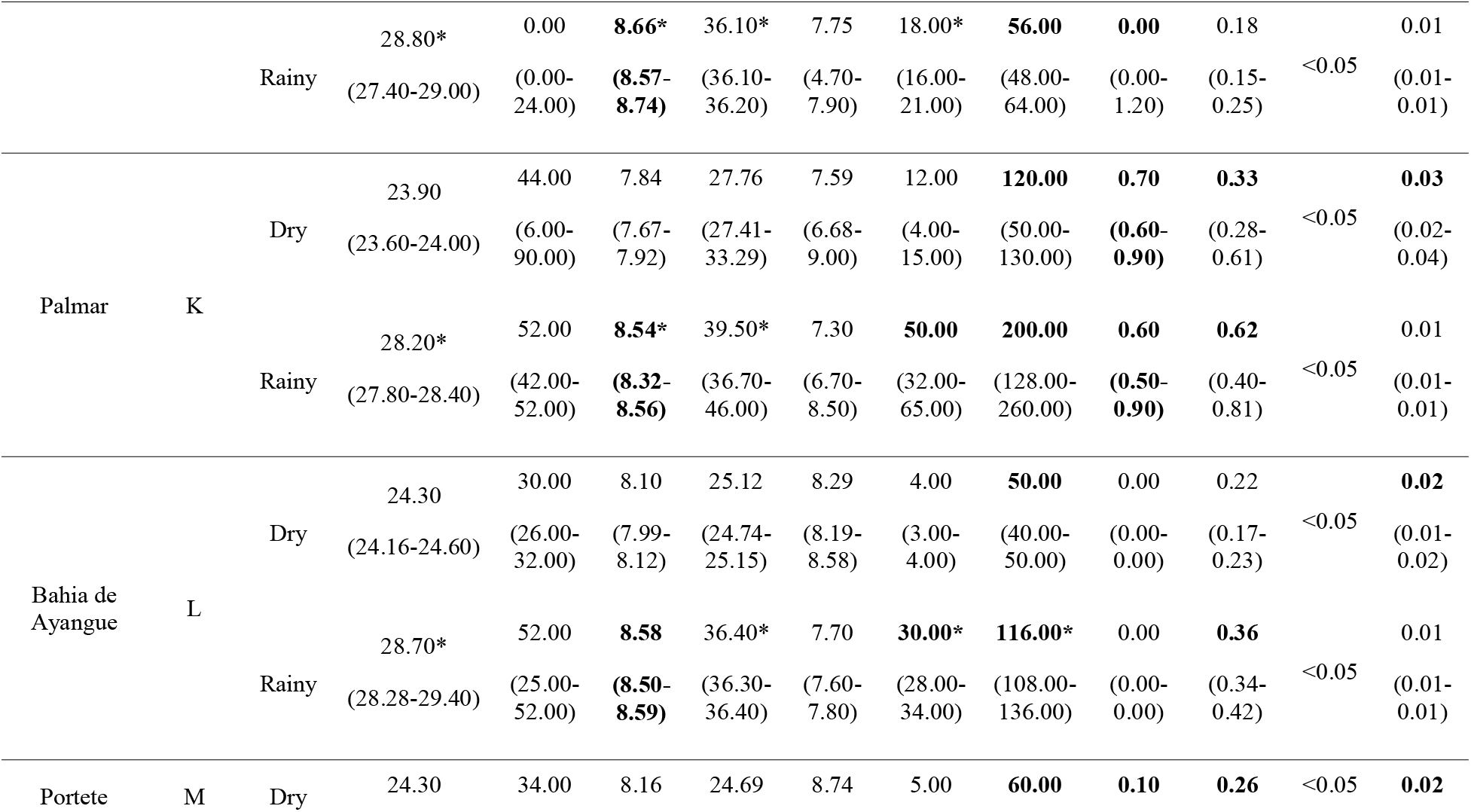
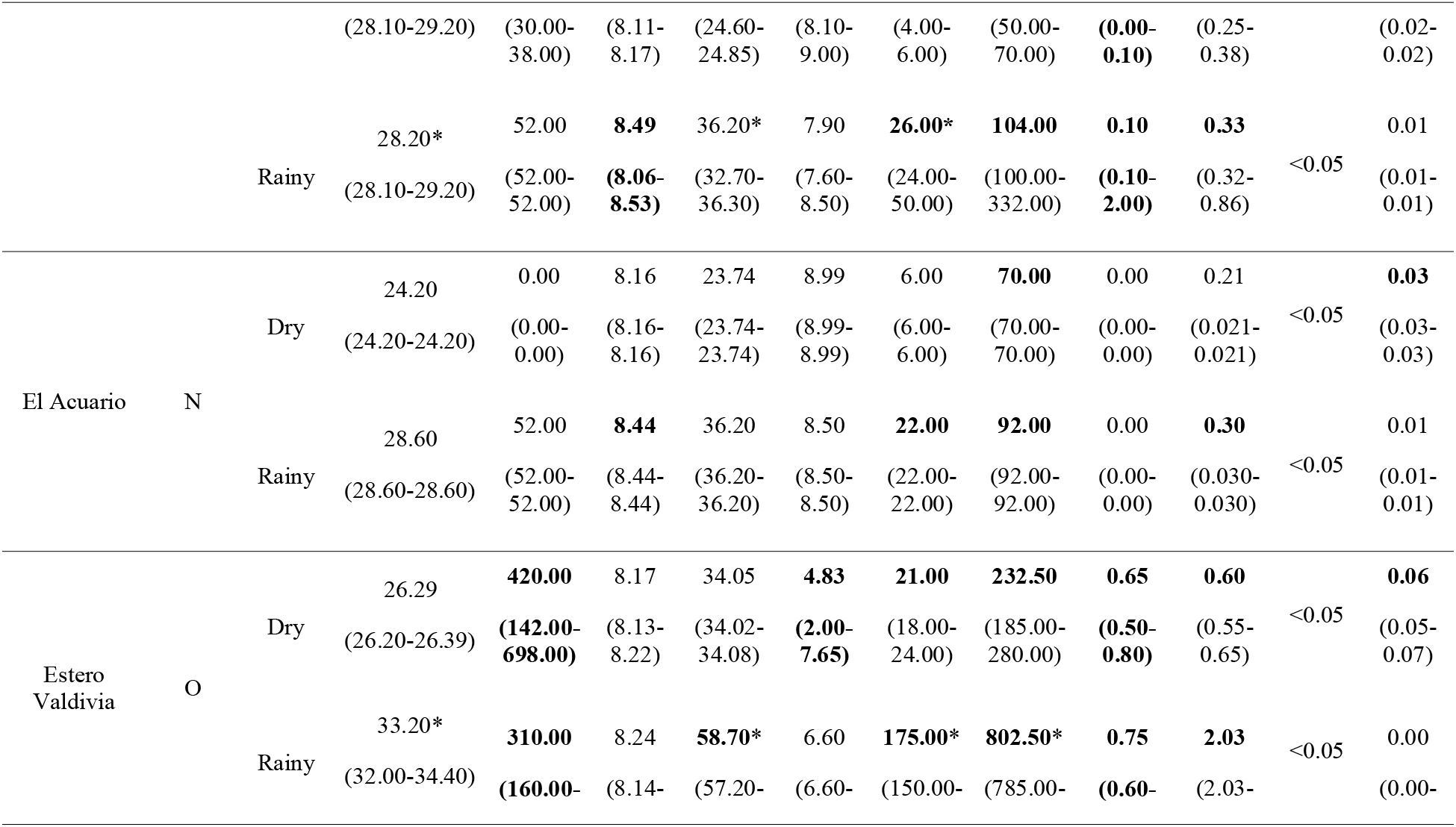
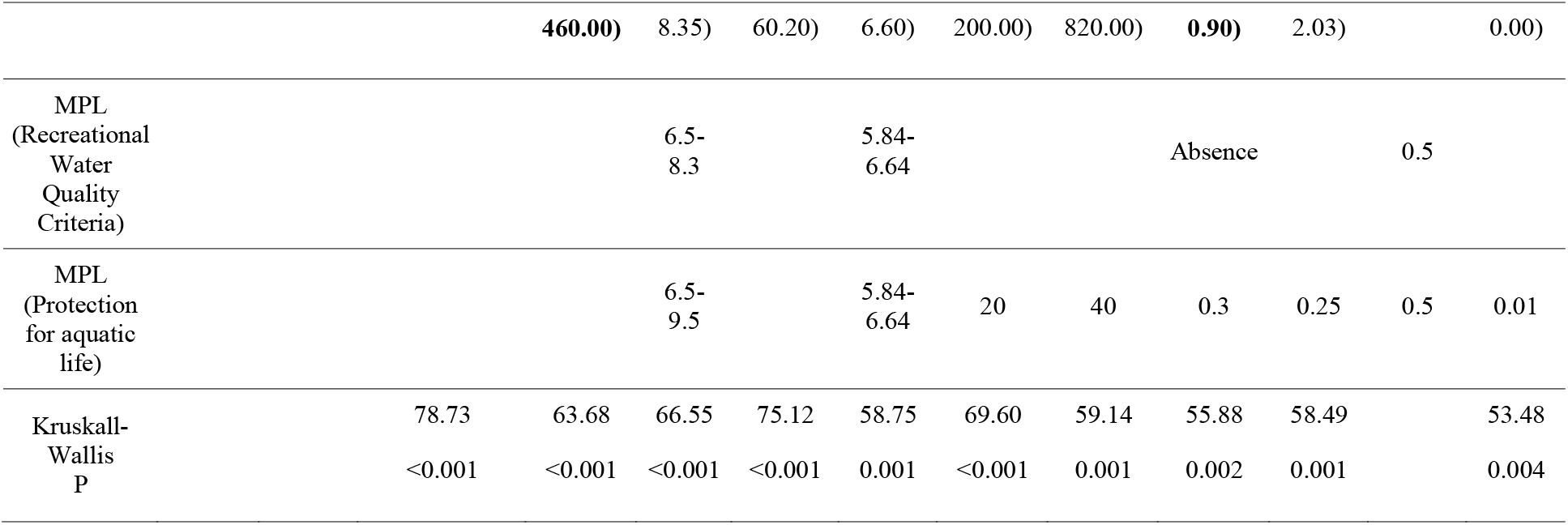
Physicochemical parameters in seawater of the beaches under study. * Indicates that there are significant differences between season in the same station according to U Mann-Whitney (P>0.05), MPL: maximum permissible level according to AM 097A. OD based on the criterion of minimum percentage of oxygen saturation 80% where the optimal values would be 7.3-8.3 OD mg / L at 25-30 ° C and 80% saturation = 5.84-6.64 mg / L.

