## Supplementary Table S1: Physicochemical parameters in seawater of the beaches under study. for "Detection of fecal coliforms and SARS-CoV-2 RNA in sewage and recreational waters in the Ecuadorian Coast: a call for improving water quality regulation"

### 3 Supplementary material

4 **Table S1.** Physicochemical parameters in seawater of the beaches under study.
5 * Indicates that there are significant differences between season in the same station according to U Mann-Whitney (P>0.05), MPL:
6 maximum permissible level according to AM 097A. OD based on the criterion of minimum percentage of oxygen saturation 80% where
7 the optimal values would be 7.3-8.3 OD mg / L at 25-30 ° C and 80% saturation = 5.84-6.64 mg / L.

### Stations Station Season Temperature

| **(°C) (mg/L)** | **(UPS)** | **(mg/L)** | **(mg/L)** | **(mg/L)** | **(mg/L)** | **(mg/L)** | **(mg/L)** | **(mg/L)** |
| --- | --- | --- | --- | --- | --- | --- | --- | --- |
| 24.20 24.00 7.98 | 22.93 | 8.12 | 5.00 | **80.00** | 0.00 | **0.45** |  | 0.02 |

**TDS**

### pH Salinity DO

**BOD**

**COD**

**O&F**

**NH_4_**

### TENS Cl

Rompeolas	A

Dry

(24.10-24.30)

30.00*

| (23.00- | (7.47- | (21.90- | (8.04- | (2.00- | (60.00- | (0.00- | (0.25- <0.05 (0.01- | |
| --- | --- | --- | --- | --- | --- | --- | --- | --- |
| 32.00) | 8.04) | 23.53) | 8.14) | 7.00) | 170.00) | 0.10) | 0.60) | 0.05) |
| 6.00 | **8.53*** | **34.30*** | 7.35 | 17.00* | **52.00** | 0.00 | **0.30** | 0.01 |

| (2.00- | (7.20- | (0.72- | (4.23- | (5.00- | (27.00- | (0.00- | (0.29- | <0.05 | (0.01- |
| --- | --- | --- | --- | --- | --- | --- | --- | --- | --- |
| 26.00) | 7.56) | 23.36) | 8.13) | 15.00) | 50.00) | 0.30) | 0.40) |  | 0.03) |
| 84.00 | **8.37*** | 0.90 | 5.83 | 44.00* | **95.00*** | **1.80** | **1.99** | <0.05 | 0.01 |

| Rainy (4.00- | (8.22- | (34.20- | (7.33- | (10.00- | (30.00- | (0.00- | (0.21- <0.05 (0.01- | |
| --- | --- | --- | --- | --- | --- | --- | --- | --- |
| (28.80-30.50) 8.00) | 8.57) | 34.30) | 7.86) | 18.00) | 56.00) | 0.00) | 0.37) | 0.01) |
| 24.70 12.00 | 7.54 | 1.17 | 5.94 | 10.00 | 30.00 | **0.20** | **0.37** | 0.01 |

Área de pescadores	B

Dry

(24.50-25.30)

Rainy	**27.30***

### Stations Station Season Temperature

| **(°C) (mg/L)** | **(UPS)** | **(mg/L)** | **(mg/L)** | **(mg/L)** | **(mg/L)** | **(mg/L)** | **(mg/L)** | **(mg/L)** |
| --- | --- | --- | --- | --- | --- | --- | --- | --- |
| (27.10-29.80) (2.00- (8.06- | (0.50- | (2.00- | (21.00- | (64.00- | (0.30- | (0.34- |  | (0.01- |
| 114.00) 8.56) | 34.50) | 7.47) | 46.00) | 96.00) | 2.00) | 8.00) |  | 0.01) |
| 25.70 26.00 7.92 | 23.57 | 7.79 | 15.00 | **130.00** | 0.00 | **0.92** |  | 0.07 |

**TDS**

### pH Salinity DO

**BOD**

**COD**

**O&F**

**NH_4_**

### TENS Cl

Parque Infantil

Dry

C

(25.60-26.00)

# 30.00*

| (22.00- | (7.82- | (23.33- | (7.72- | (12.00- | (120.00- | (0.00- | (0.89- <0.05 (0.06- | |
| --- | --- | --- | --- | --- | --- | --- | --- | --- |
| 28.00) | 8.09) | 23.60) | 7.92) | 20.00) | 180.00) | 0.00) | 0.98) | 0.08) |
| 4.00 | **8.66*** | 34.30* | 7.45* | 32.00 | **100.00** | 0.00 | **0.38** | 0.01 |

| Rainy (4.00- | (8.30- | (34.30- | (7.42- | (12.00- | (40.00- | (0.00- | (0.24- <0.05 (0.01- | |
| --- | --- | --- | --- | --- | --- | --- | --- | --- |
| (29.60-30.20) 6.00) | 8.69) | 34.80) | 7.55) | 35.00) | 104.00) | 0.00) | 0.39) | 0.01) |
| 25.70 32.00 | 8.29 | 5.80 | 9.00 | 7.00 | **80.00** | **0.30** | **1.06** | 0.03 |

Esterillo	D

Dry

(25.10-26.20)

# 31.60*

| (22.00- | (7.68- | (5.47- | (7.84- | (5.00- | (60.00- | (0.10- | (0.25- <0.05 (0.02- | |
| --- | --- | --- | --- | --- | --- | --- | --- | --- |
| 36.00) | 8.45) | 23.37) | 9.00) | 10.00) | 100.00) | 1.80) | 4.70) | 0.04) |
| 50.00 | 8.07 | 0.90 | **3.47** | 11.00 | **50.00** | **1.10** | **1.93** | 0.01 |

| Rainy (42.00- | (7.72- | (0.80- | (3.10- | (8.4- | (34.00- | (0.50- | (0.16- | <0.05 | (0.01- |
| --- | --- | --- | --- | --- | --- | --- | --- | --- | --- |
| (31.40-33.40) 86.00) | 8.42) | 16.50) | 5.20) | 42.00) | 89.00) | 1.70) | 2.14) |  | 0.01) |
| 26.60  Estero Data E Dry 24.00 | 7.10 | 22.43 | 7.67 | 8.00 | **90.00** | **1.00** | 0.15 | <0.05 | **0.04** |

(25.40-27.20)

### Stations Station Season Temperature

| **(°C) (mg/L)** | **(UPS)** | **(mg/L)** | **(mg/L)** | **(mg/L)** | **(mg/L)** | **(mg/L)** | **(mg/L)** | **(mg/L)** |
| --- | --- | --- | --- | --- | --- | --- | --- | --- |
| (14.00- | (6.88- (21.28- | (7.07- | (6.00- | (70.00- | (0.10- | (0.12- |  | **(0.03-** |
| 34.00) | 8.36) 27.16) | 9.00) | 8.00) | 90.00) | 1.90) | 0.19) |  | **0.04)** |
| 31.30* 38.00 | 7.92 34.30* | 5.70* | **76.00** | **302.00** | **1.00** | **0.78** |  | 0.10* |
| Rainy (4.00- | (7.73- (21.28- | (4.59- | (32.00- | (96.00- | (0.07- | (0.36- | <0.05 (0.01- | |
| (31.00-31.30) 52.00) | 8.29) 27.16) | 7.05) | 80.00) | 384.00) | 2.00) | 0.98) | 0.10) | |
| 24.00 10.00 | 7.28 25.85 | 5.21 | 8.00 | **90.00** | **0.50** | 0.14 | **0.03** | |

**TDS**

### pH Salinity DO

**BOD**

**COD**

**O&F**

**NH_4_**

### TENS Cl

Facilidad Pesquera	F Anconcito

Dry

(23.90-24.10)

27.00*

| (8.00- | (7.20- | (25.40- | (2.98- | (6.00- | (70.00- | (0.50- | (0.13- <0.05 (0.03- | |
| --- | --- | --- | --- | --- | --- | --- | --- | --- |
| 14.00) | 7.30) | 25.94) | 6.17) | 18.00) | 160.00) | 0.60) | 0.16) | 0.070) |
| 42.00 | **8.53*** | 36.00* | 6.92* | **30.00** | **92.00** | **2.00** | **0.35** | 0.10 |

| Rainy (6.00- | (7.93- | (35.90- | (3.21- | (25.00- | (76.00- | (0.40- | (0.31- <0.05 (0.01- | |
| --- | --- | --- | --- | --- | --- | --- | --- | --- |
| (26.70-27.60) 52.00) | 8.64) | 36.20) | 7.05) | 38.00) | 116.00) | 2.60) | 0.41) | 0.10) |
| 24.50 26.00 | 7.30 | 24.99 | 8.16 | 5.00 | **60.00** | 0.00 | 0.17 | **0.02** |

Punta Carnero

Dry

G

Rainy

(24.40-24.70)

26.90* (26.60-27.30)

| (22.00- | (7.17- | (20.14- | (7.58- | (4.00- | (84.00- | (0.00- | (0.15- | <0.05 | (0.01- |
| --- | --- | --- | --- | --- | --- | --- | --- | --- | --- |
| 28.00) | 7.62) | 25.36) | 8.16) | 30.00) | 96.00) | 0.70) | 5.35) |  | 0.08) |
| 2.00 | **8.48*** | 36.50* | 7.80 | **29.00** | **88.00** | **0.40** | **0.34** | <0.05 | 0.01 |

### Stations Station Season Temperature

| **(°C) (mg/L)** | **(UPS)** | **(mg/L)** | **(mg/L)** | **(mg/L)** | **(mg/L)** | **(mg/L)** | **(mg/L)** | **(mg/L)** |
| --- | --- | --- | --- | --- | --- | --- | --- | --- |
| (2.00- | (8.47- (35.90- | (7.62- | (28.00- | (84.00- | (0.00- | (0.32- |  | (0.01- |
| 6.00) | 8.67) 36.70) | 7.85) | 32.00) | 96.00) | 0.40) | 0.38) |  | 0.01) |
| 24.70 24.00 | 7.26 25.25 | 8.03 | 8.00 | **90.00** | **0.10** | 0.13 |  | **0.03** |

**TDS**

### pH Salinity DO

**BOD**

**COD**

**O&F**

**NH_4_**

### TENS Cl

Chipipe	H

Dry

(24.50-24.70)

28.40*

| (22.00- | (7.00- | (24.96- | (7.84- | (7.00- | (80.00- | (0.10- | (0.13- <0.05 (0.02- | |
| --- | --- | --- | --- | --- | --- | --- | --- | --- |
| 26.00) | 7.86) | 25.26) | 8.04) | 10.00) | 100) | 0.10) | 0.19) | 0.03) |
| 0.00 | **8.59*** | 36.00 | 7.55* | **32.00*** | **96.00*** | **0.10** | **0.36** | 0.01 |

| Rainy (0.00- | (8.34- | (36.00- | (7.55- | (30.00- | (88.00- | (0.00- | (0.33- <0.05 (0.01- | |
| --- | --- | --- | --- | --- | --- | --- | --- | --- |
| (27.90-28.50) 0.00) | 8.75) | 36.10) | 7.64) | 34.00) | 100.00) | 0.10) | 0.41) | 0.01) |
| 24.40 28.00 | 7.40 | 25.55 | 7.83 | 4.00 | **50.00** | **0.10** | 0.14 | **0.02** |

San Lorenzo	I

Dry

(24.20-27.70)

27.60

| (26.00- | (7.15- | (25.22- | (7.70- | (3.00- | (40.00- | (0.00- | (0.12- <0.05 (0.01- | |
| --- | --- | --- | --- | --- | --- | --- | --- | --- |
| 30.00) | 7.46) | 25.76) | 8.45) | 4.00) | 50.00) | 0.20) | 0.15) | 0.02) |
| 0.00 | **8.69*** | 36.00* | 7.61* | **25.00*** | **76.00*** | **0.10** | **0.31** | 0.01 |

| Rainy (0.00- | **(8.68-** | (36.00- | (7.44- | (22.00- | (68.00- | (0.10- | (0.28- | <0.05 | (0.01- |
| --- | --- | --- | --- | --- | --- | --- | --- | --- | --- |
| (27.30-28.00) 0.00) | **8.73)** | 36.10) | 7.64) | 28.00) | 88.00) | 0.30) | 0.34) |  | 0.01) |
| 24.50  Santa Rosa J Dry 26.00 | 7.32 | 25.60 | 8.91 | 8.00 | **90.00** | **0.30** | 0.18 | <0.05 | **0.03** |

(24.40-24.60)

### Stations Station Season Temperature

| **(°C) (mg/L)** | **(UPS)** | **(mg/L)** | **(mg/L)** | **(mg/L)** | **(mg/L)** | **(mg/L)** | **(mg/L)** | **(mg/L)** |
| --- | --- | --- | --- | --- | --- | --- | --- | --- |
| (22.00- | (7.25- (25.33- | (8.11- | (3.00- | (40.00- | (0.20- | (0.16- |  | (0.01- |
| 28.00) | 8.29) 25.90) | 9.00) | .00) | 90.00) | 0.50) | 0.20) |  | 0.03) |
| 28.80* 0.00 | **8.66*** 36.10* | 7.75 | 18.00* | **56.00** | **0.00** | 0.18 |  | 0.01 |
| Rainy (0.00- | **(8.57-** (36.10- | (4.70- | (16.00- | (48.00- | (0.00- | (0.15- | <0.05 (0.01- | |
| (27.40-29.00) 24.00) | **8.74)** 36.20) | 7.90) | 21.00) | 64.00) | 1.20) | 0.25) | 0.01) | |
| 23.90 44.00 | 7.84 27.76 | 7.59 | 12.00 | **120.00** | **0.70** | **0.33** | **0.03** | |

**TDS**

### pH Salinity DO

**BOD**

**COD**

**O&F**

**NH_4_**

### TENS Cl

Palmar	K

Dry

(23.60-24.00)

28.20*

| (6.00- | (7.67- | (27.41- | (6.68- | (4.00- | (50.00- | **(0.60-** | (0.28- <0.05 (0.02- | |
| --- | --- | --- | --- | --- | --- | --- | --- | --- |
| 90.00) | 7.92) | 33.29) | 9.00) | 15.00) | 130.00) | **0.90)** | 0.61) | 0.04) |
| 52.00 | **8.54*** | 39.50* | 7.30 | **50.00** | **200.00** | **0.60** | **0.62** | 0.01 |

| Rainy (42.00- | **(8.32-** | (36.70- | (6.70- | (32.00- | (128.00- | **(0.50-** | (0.40- <0.05 (0.01- | |
| --- | --- | --- | --- | --- | --- | --- | --- | --- |
| (27.80-28.40) 52.00) | **8.56)** | 46.00) | 8.50) | 65.00) | 260.00) | **0.90)** | 0.81) | 0.01) |
| 24.30 30.00 | 8.10 | 25.12 | 8.29 | 4.00 | **50.00** | 0.00 | 0.22 | **0.02** |

Bahia de Ayangue

Dry

L

Rainy

(24.16-24.60)

28.70* (28.28-29.40)

| (26.00- | (7.99- | (24.74- | (8.19- | (3.00- | (40.00- | (0.00- | (0.17- | <0.05 | (0.01- |
| --- | --- | --- | --- | --- | --- | --- | --- | --- | --- |
| 32.00) | 8.12) | 25.15) | 8.58) | 4.00) | 50.00) | 0.00) | 0.23) |  | 0.02) |
| 52.00 | **8.58** | 36.40* | 7.70 | **30.00*** | **116.00*** | 0.00 | **0.36** | <0.05 | 0.01 |

### Stations Station Season Temperature

| **(°C) (mg/L)** | **(UPS)** | **(mg/L)** | **(mg/L)** | **(mg/L)** | **(mg/L)** | **(mg/L)** | **(mg/L)** | **(mg/L)** |
| --- | --- | --- | --- | --- | --- | --- | --- | --- |
| (25.00- | **(8.50-** (36.30- | (7.60- | (28.00- | (108.00- | (0.00- | (0.34- |  | (0.01- |
| 52.00) | **8.59)** 36.40) | 7.80) | 34.00) | 136.00) | 0.00) | 0.42) |  | 0.01) |
| 24.30 34.00 | 8.16 24.69 | 8.74 | 5.00 | **60.00** | **0.10** | **0.26** |  | **0.02** |

**TDS**

### pH Salinity DO

**BOD**

**COD**

**O&F**

**NH_4_**

### TENS Cl

Portete	M

Dry

(28.10-29.20)

28.20*

| (30.00- | (8.11- | (24.60- | (8.10- | (4.00- | (50.00- | **(0.00-** | (0.25- <0.05 (0.02- | |
| --- | --- | --- | --- | --- | --- | --- | --- | --- |
| 38.00) | 8.17) | 24.85) | 9.00) | 6.00) | 70.00) | **0.10)** | 0.38) | 0.02) |
| 52.00 | **8.49** | 36.20* | 7.90 | **26.00*** | **104.00** | **0.10** | **0.33** | 0.01 |

| Rainy (52.00- | **(8.06-** | (32.70- | (7.60- | (24.00- | (100.00- | **(0.10-** | (0.32- <0.05 (0.01- | |
| --- | --- | --- | --- | --- | --- | --- | --- | --- |
| (28.10-29.20) 52.00) | **8.53)** | 36.30) | 8.50) | 50.00) | 332.00) | **2.00)** | 0.86) | 0.01) |
| 24.20 0.00 | 8.16 | 23.74 | 8.99 | 6.00 | **70.00** | 0.00 | 0.21 | **0.03** |

El Acuario	N

Dry

(24.20-24.20)

28.60

| (0.00- | (8.16- | (23.74- | (8.99- | (6.00- | (70.00- | (0.00- | (0.021- <0.0 | ^5^ (0.03- |
| --- | --- | --- | --- | --- | --- | --- | --- | --- |
| 0.00) | 8.16) | 23.74) | 8.99) | 6.00) | 70.00) | 0.00) | 0.021) | 0.03) |
| 52.00 | **8.44** | 36.20 | 8.50 | **22.00** | **92.00** | 0.00 | **0.30** | 0.01 |

| Rainy (52.00- | (8.44- | (36.20- | (8.50- | (22.00- | (92.00- | (0.00- | (0.030- | <0.05 | (0.01- |
| --- | --- | --- | --- | --- | --- | --- | --- | --- | --- |
| (28.60-28.60) 52.00) | 8.44) | 36.20) | 8.50) | 22.00) | 92.00) | 0.00) | 0.030) |  | 0.01) |
| 26.29  Estero O Dry **420.00** | 8.17 | 34.05 | **4.83** | **21.00** | **232.50** | **0.65** | **0.60** | <0.05 | **0.06** |

Valdivia	(26.20-26.39)

### Stations Station Season Temperature

| **(°C) (mg/L)** | **(UPS)** | **(mg/L)** | **(mg/L)** | **(mg/L)** | **(mg/L)** | **(mg/L)** | **(mg/L)** | **(mg/L)** |
| --- | --- | --- | --- | --- | --- | --- | --- | --- |
| **(142.00-** (8.13- | (34.02- | **(2.00-** | (18.00- | (185.00- | **(0.50-** | (0.55- |  | (0.05- |
| **698.00)** 8.22) | 34.08) | **7.65)** | 24.00) | 280.00) | **0.80)** | 0.65) |  | 0.07) |
| 33.20* **310.00** 8.24 | **58.70*** | 6.60 | **175.00*** | **802.50*** | **0.75** | **2.03** |  | 0.00 |

**TDS**

### pH Salinity DO

**BOD**

**COD**

**O&F**

**NH_4_**

### TENS Cl

Rainy

(32.00-34.40)

MPL

| **(160.00-** | (8.14- | (57.20- | (6.60- | (150.00- | (785.00- | **(0.60-** | (2.03- <0.05 (0.00- | |
| --- | --- | --- | --- | --- | --- | --- | --- | --- |
| **460.00)** | 8.35) | 60.20) | 6.60) | 200.00) | 820.00) | **0.90)** | 2.03) | 0.00) |

(Recreational Water Quality Criteria)

MPL

6.5-

8.3

6.5-

5.84-

6.64

5.84-

Absence	0.5

(Protection for aquatic life)

9.5

6.64	20	40	0.3	0.25	0.5	0.01

Kruskall- Wallis

P

78.73

<0.001

63.68

<0.001

66.55

<0.001

75.12

<0.001

58.75

0.001

69.60

<0.001

59.14

0.001

55.88

0.002

58.49

0.001

53.48

0.004

8
